# Simplified model of Covid-19 epidemic prognosis under quarantine and estimation of quarantine effectiveness

**DOI:** 10.1101/2020.04.28.20083428

**Authors:** Algis Džiugys, Martynas Bieliūnas, Gediminas Skarbalius, Edagaras Misiulis, Robertas Navakas

## Abstract

A simplified model of Covid-19 epidemic dynamics under quarantine conditions and method to estimate quarantine effectiveness are developed. The model is based on the growth rate of new infections when total number of infections is significantly smaller than population size of infected country or region. The model is developed on the basis of collected epidemiological data of Covid19 pandemic, which shows that the growth rate of new infections has tendency to decrease linearly when the quarantine is imposed in a country (or a region) until it reaches a constant value, which corresponds to the effectiveness of quarantine measures taken in the country. The growth rate of new infections can be used as criteria to estimate quarantine effectiveness.

## Introduction

The 2019–2020 coronavirus outbreak has started since December 29th, 2019 in Wuhan, Hubei province, People’s Republic of China, and has progressively expanded through almost all countries. This ongoing pandemic of coronavirus disease 2019 (COVID-19) caused by severe acute respiratory syndrome coronavirus 2 (SARS-CoV-2).

In order to prevent the spread of the disease, many of the countries affected by the disease has been put under quarantine, which has led that 4,5 billion people (58 % of whole world population) around the globe faced some form of lockdown [1]. However, the remaining problem of the current COVID-19 disease is that second wave of the epidemic is expected after imposed quarantine is discontinued in each country. On the basis of epidemiological data provided by European Centre for Disease Prevention and Control [2] and estimation that the number of infections can be four times higher than the number of confirmed cases [3], it can be evaluated that for most countries only less than 10 % of the population of the infected country/region will acquire the immunity after first wave of epidemic controlled by the effective quarantine. That would require next 5-6 similarly controlled waves to achieve herd (population) immunity of 50-70 % in order to stop further spread of disease. Such scenario is improbable as it would require a significant amount of time, during which the individual immunity could be lost because even today, when more than 4.8 million people have been infected in more than 188 countries and territories (20 May 2020, [4]), it is not clear for how long time the recovered patients have the immunity. However, hope for SARS-CoV-2 antiviral vaccine or sufficiently effective antiviral drugs, or SARS-CoV-2 virus mutation to less aggressive strain [5] gives us chance to survive through the course of the controlled epidemic.

Therefore, forecasting the spread of the pandemic on the basis of mathematical models are extremely important for decisions how to prepare countries in order to avoid overloading of health system and manage other related problems. Valuable information that could be obtained from modelling is forecast of the expected time and number of most active infected cases and the effectiveness of applied infection control measures. It is current global trend, that the experience and available data from already affected countries are used to model the pandemic dynamics in other countries before the epidemic has reached the peak or to estimate effectiveness of various scenario of the next wave management [6].

Most popular epidemic dynamics models of Covid-19 are based on transmission model for a directly transmitted infectious disease, such as standard compartment models of disease SIR [7,8], or more advances derivates, such as SEIR and similar models [9–12]. Many of the models, which are used to forecast the COVID-19 epidemic, do not accurately capture the transient dynamics of epidemics; therefore, they give poor predictions of both the epidemic’s peak and its duration [13], because calibration of parameters are based on dynamics of such non-reliable epidemiological data as number of active infectious cases.

We propose to build epidemic analysis and model on the dynamics of rate of new infection cases as more reliable epidemiological data together with an assumption of effectiveness to isolate registered infectious during imposed quarantine. Other epidemiological parameters, such as numbers of active infectious, deaths, recovered, severe patients and others, can be forecasted from the forecasted dynamics of the rate of new infectious cases. The proposed approach is based on SIR model.

### SIR model

The simplest SIR model consists of three compartments: *S* for the number of susceptible, *I* for the number of infectious, and *R* for the number of removed (recovered, deceased or immune) individuals. These variables (*S, I*, and *R*) represent the number of people in each compartment at a particular time. We denote the total population size by *N*. The dynamics of the simplest SIR system (excluding birth and death) can be described by the following set of ordinary differential equations [11]:

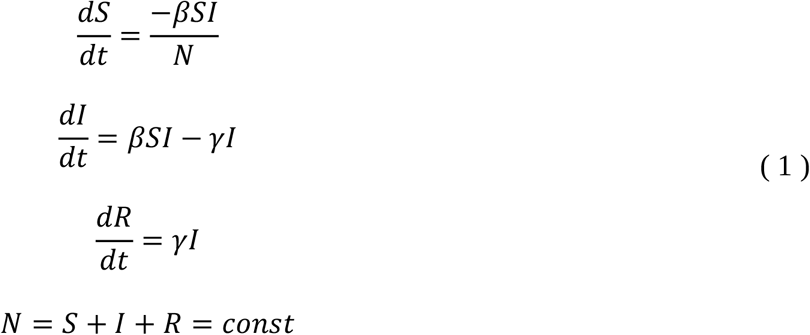

where *γ* is the rate of recovery or mortality, *β* is the infectious rate controlling the rate of spread that represents the probability of transmitting disease from infectious individual to susceptible individual. The disease transition rate *β/NI* is defined as a product of *β* and a probability of disease transmission during a contact between an infectious individual and a susceptible individual. In that case, *β* is the average number of contacts per person per time unit and can be defined by the typical time between contacts *T_c_ =* 1/*β*. The transition rate between *I* and *R* defined by *γ* and is estimated from typical time until recovery *T_r_ =* 1*/γ*. In case of isolation or self-isolation, *γ* can be defined by the average number of days *T_ri_* that a person is infectious (before they are isolated or self-isolate), γ = 1*/T_ri_*.

The dynamics of the infectious class depends on the following ratio:

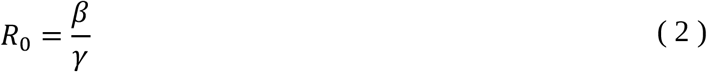

the so-called basic reproduction number (or basic reproduction ratio) of an infection and represents the average number of infections generated by one individual over the course of the infectious period.

The estimation model parameters and *β*, as *γ*, well as *R*_0_, may vary depending on country due to methodological issues, including different assumptions and choice of parameters, utilized models, used datasets and estimation period. In addition, during the spread of the SARS-CoV-2 virus infection, it was found that the model parameters are varying according to the dynamics of transmission of the novel coronavirus outbreak as well as the case reporting rate, which requires to build up more sophisticated and complex models [14].The model parameters are constantly calibrated according to the last epidemiological data because forecast is constantly refreshed.

### Simplified model of epidemic dynamics under quarantine

The wide and rapid spread of the disease forced many countries to impose quarantines, entry bans and other restrictions during the pandemic to reduce the movement of population and recent travellers in most affected regions [15]. Global restrictions that apply to all foreign countries and regions have also been imposed in other countries preventing their own citizens to travel overseas. These measures, especially quarantine, helped to suppress the spread of the disease within the population of various countries with different effectiveness.

Other important property of the pandemic is that total number of infected cases *T* is much smaller than population size of infected country or region due to quarantine measures:

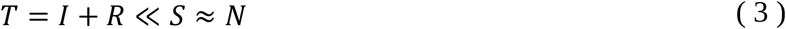

This assumption allows to simplify epidemiological models used to simulate Codiv-19 disease spread under quarantine.

Consequently, on the basis of eq. (3), the parameter *β* of SIR model can be estimated as the number of new registered cases of infection to number of active cases ratio:

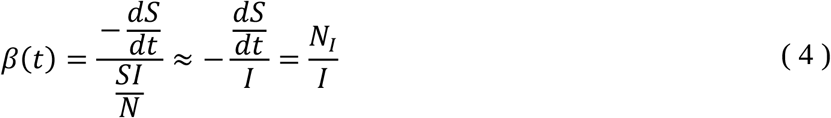

where *N_I_* is the rate of new infected cases

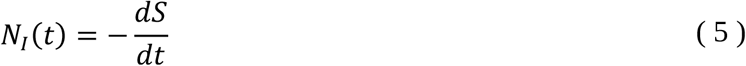

which can be estimated by counting new cases of infection, and usually is measured by number of registered new cases per time period *τ*

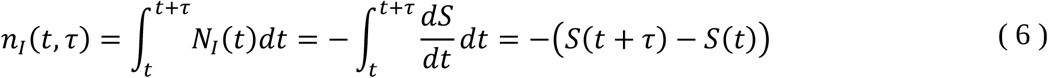

The number of daily new infected cases is defined as

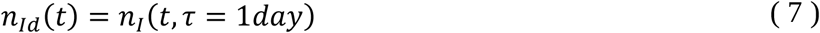

So, expected number of new cases in next day could be predicted by the today number of active infected individuals multiplied by the infectious rate

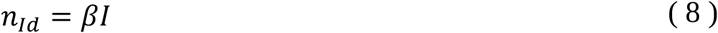

In general, the infectious rate *β* is time dependent and usually can be described by a complex function with additional parameters that must be daily calibrated according to last epidemiological data. Behaviour of the infectious rate *β* during quarantine may differ in different countries, which reduces possibilities to build correct model of the infectious rate on the basis of epidemiological data from other countries. Furthermore, there is no clear criteria to relate influence of quarantine actions with the infectious rate *β*.

As illustration, the dynamics of the parameter *β* estimated from active infected individuals and new cases by eq. (4) is demonstrated in Figure 1 for 46 countries (Albania, Algeria, Argentina, Australia, Austria, Azerbaijan, Bangladesh, Belarus, Belgium, Bosnia and Herzegovina, Chile, Croatia, Denmark, Ecuador, Estonia, Finland, Germany, Greece, Hungary, Iceland, Ireland, Italy, Kazakhstan, Latvia, Lithuania, Luxembourg, Malaysia, Malta, Moldova, Montenegro, Netherlands, New Zealand, Norway, Poland, Portugal, Romania, Russia, San Marino, Serbia, Slovakia, Spain, Sweden, Switzerland, Tunisia, Ukraine, United Kingdom) with imposed quarantine. The applied control measures of the quarantine in each country are described in [16]. Time is aligned to the date of imposed quarantine in each country *t*′ = *t* − *t_q_*, where *t_q_* is the initialisation date of the quarantine [16]. Daily numbers of active infected individuals and new infection cases until 20 May 2020 were collected from open sources [17]. In order to damp daily and weekly fluctuations, *I*, *n_Id_* and estimated the parameter *β* were smoothed by moving average, where each average is calculated over a sliding window of length *N_s_* = 7 days:

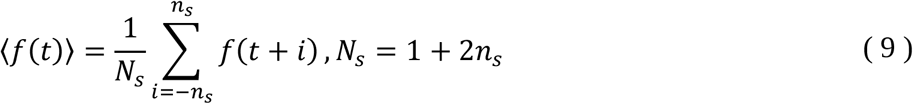

where *f* is any parameter to be smoothed.

As can be seen, the dynamics of the parameter *β* demonstrates similar non-linearly decreasing behaviour after the start of the quarantine for almost all countries.

**Figure 1.**
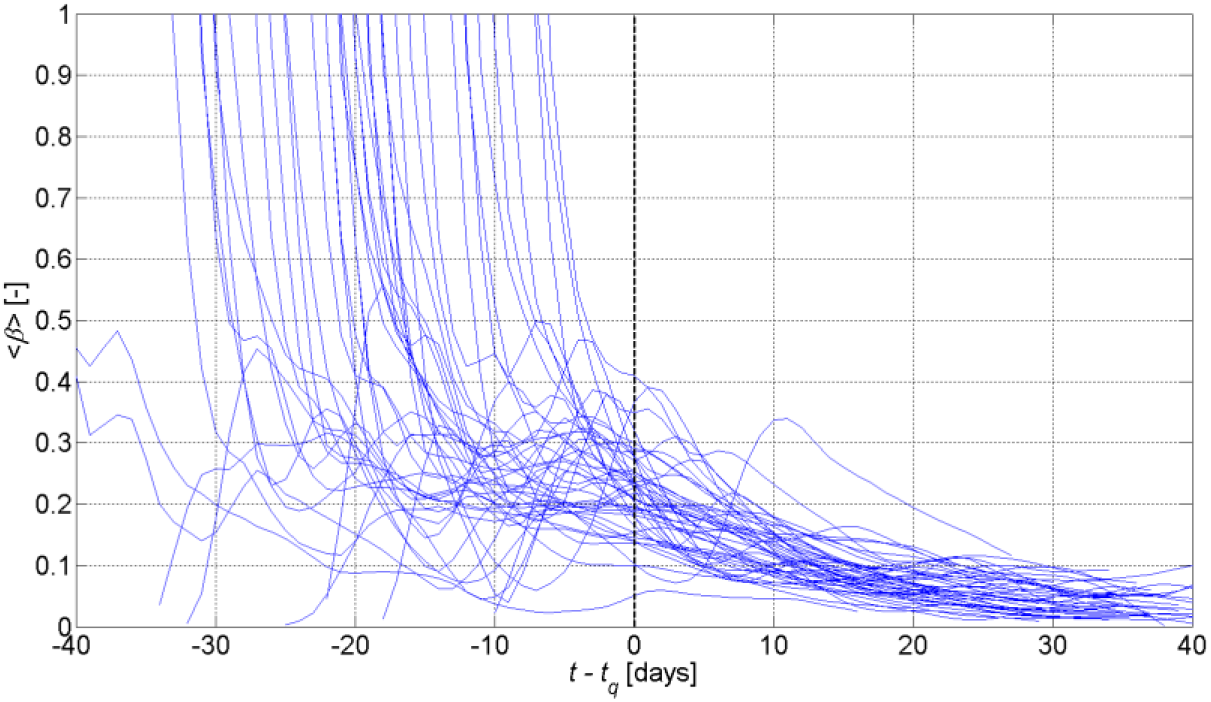
The averaged infectious rate 〈*β*〉 versus time for 46 countries. Vertical black line points the day of imposed quarantine.

In addition, an estimation of the number of active infected individuals *I* depends on the number of recovered active cases of infections *R*, which in turn depends on testing protocols, tests number, delays in testing and other circumstances, which are different in each country. Consequently, this makes *I = T − R* unreliable parameter for model calibration. For example, in Lithuania the first pool consisting of 54 recovered cases of 999 total cases was reported only after 30 days from the first infection registration [18], which can be explained only by troubles in recording of recovered cases and can be also expected in other countries. In contrary, the number of daily new infected cases, *n_Id_*, despite of countries specificity, is still the most reliable parameter allowing to estimate disease spreading. In addition, other epidemiological parameters, such as numbers of active infectious, deaths, recovered, severe patients and others, can be forecasted from the forecasted dynamics of the number of daily new infectious cases.

In order to build model of Covid-19 disease spread during quarantine, we make simplifying assumptions, that:

- registered infected individuals do not spread virus to health individuals, because of an effective isolation of the registered infected individuals;
- no imported infections.

New cases of infectious individuals are generated by previously infectious individuals until they are registered; therefore, the number of new cases of the day is dependent on the number of new cases generated during previous day and the effectiveness of imposed quarantine actions: 1) restriction of social contacts and mobility, 2) identification of infectious individual and his contact tracing as soon as possible, 3) isolation of infectious individuals.

Let us analyse hypothetical simplified case. The averaged time period, during which an infected individual is registered and isolated after infection, is denoted *T_i_*. During this time period, the infected individual is infectious and infects *α* individuals. In the end of this period, the infected individual is isolated and does not take more participation in the process of the epidemic spread. During the time period [*t* − *T_i_,t*], new infected cases are generated, from which *n_I_*(*t* − *T_i_,T_i_*) are registered and isolated. These new registered infected cases will then generate more cases and the number of new registered infection cases for next time period [*t*, *t* + *T_i_*]:

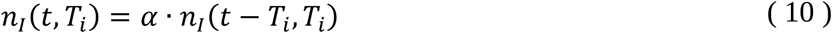

where *α* is the growth rate of new cases and can be associated with the effective reproduction number [19]. The parameter *α* is time dependent and *α* > 1 means that number of new cases is increasing, while *α* < 1 - decreasing.

According to eq. (6), the parameter *α* can expressed as follows:

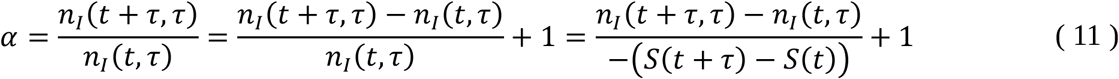

It is evident that in limits of *τ* → 0, we have

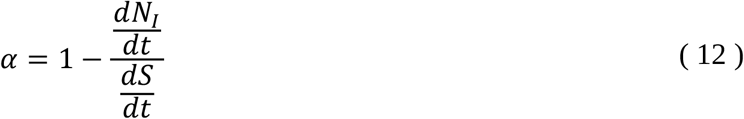

which, together with eq. (5), gives differential equation for disease dynamics during quarantine:

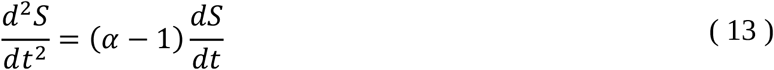

Keeping in mind eqs. (4) and (5), the parameter *α* can be related to the infectious rate *β* as follows

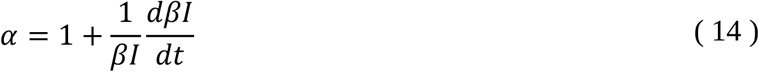

Let us define *α*_0_ as the growth rate of new infection cases when epidemic starts and spreads without control. Because of *α*_0_ > 1, the number of new cases is exponentially increasing. It is expected that the population in any country would start to behave more safely even though no official quarantine actions were taken; consequently, the growth rate of new cases is expected to slowly decrease before strict quarantine rules are imposed. The growth rate of new infected cases can be defined as *α*_*q*0_ = *α* (*t* = *t_q_*) > 1 at the quarantine start time *t* = *t_q_*. Because of *α_q_*_0_ > 1, the number of new cases is still increasing. Let us assume that the imposed quarantine is ideally effective, which means that all infected individuals are isolated until the end of the time span and do not have contact with other individuals during time span [*t_q_*, *t_q_* + *T_i_*]. Furthermore, no new cases will be generated for the next time period [*t_q_* + *T_i_*, *t_q_* + 2*T_i_*]. In such case, the growth rate of new cases becomes constant and equal to 0 after the quarantine starts: *α*(*t* > *t_q_*) = *α_q_* = 0 (Figure 2 (a)), which means that the supposed effectiveness of the quarantine is equal to 1 and epidemic is stopped immediately.

**Figure 2.**
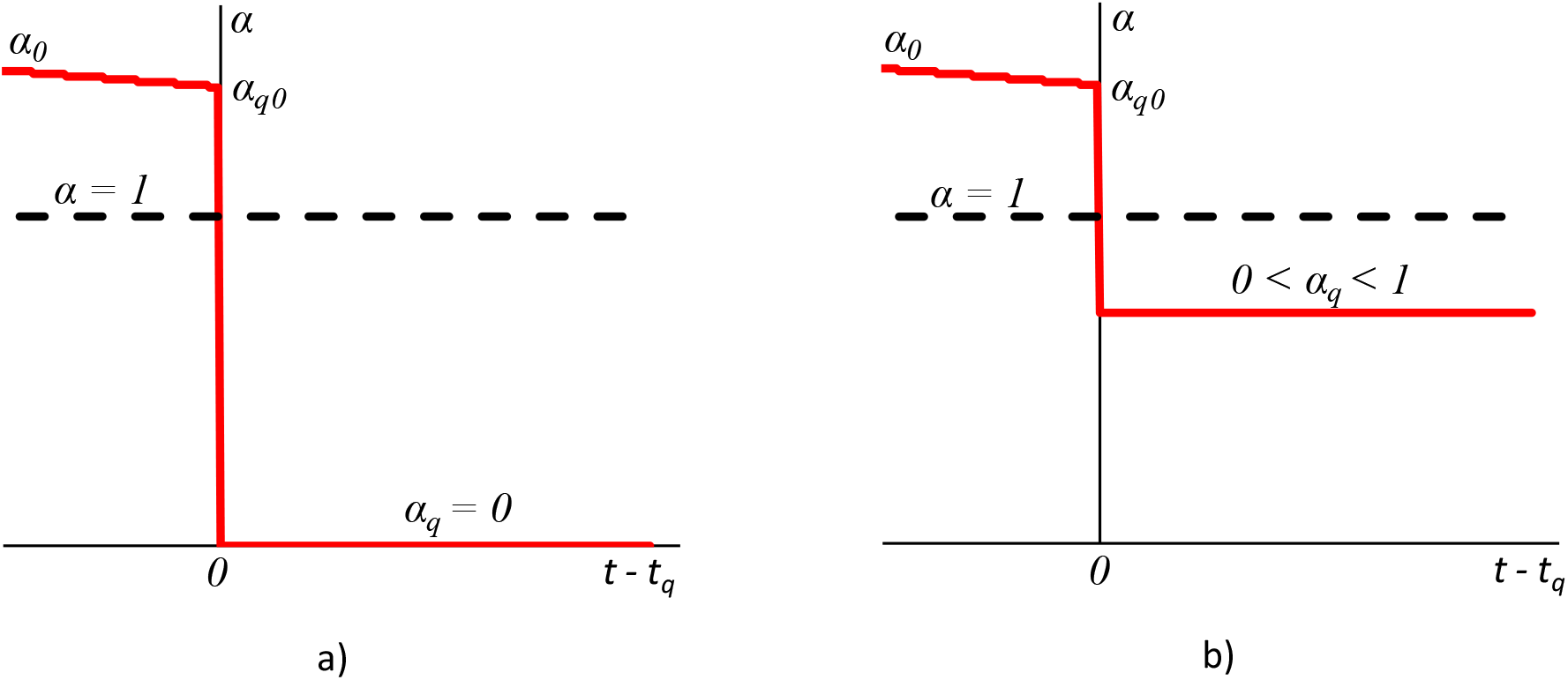
Idealized scenario of the growth rate of new cases *α* dynamics with quarantine effectiveness *e_q_*: a) *e_q_* = 1, b) *e_q_*< 1.

If the quarantine is less effective, then the growth rate of new cases is non zero constant *α*(*t* > *t_q_*) = *α_q_* > 0, which leads to slower spread of the disease in case of *α_q_*_0_ > *α_q_ >* 1 and suppression of the epidemic in case of 1 > *α_q_* > 0 (Figure 2 (b)). Consequently, quarantine effectiveness can be measured as

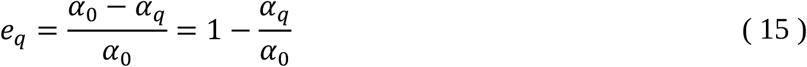

The zero effectiveness *e_q_* = 0 means that the growth rate of new cases during quarantine remains the same as before: *α_q_* = *α_q_*_0_. Therefore, in order to suppress the disease, the growth rate of new cases must be below 1, which means that quarantine effectiveness *e_q_* must be greater than 1 − 1/*α*_0_.

In realistic scenario, the growth rate of new cases *α* does not change sharply at the time *t_q_* because of time lag due to incubation period, infections generated by non-registered infected individuals and so on. In addition, it takes a time for people to adjust to the quarantine requirements after the beginning of the quarantine; therefore, there is a time lag before people start to strictly follow the rules. Consequently, the growth rate of new cases *α* decreases from initial value *α_q_*_0_ until reaches constant value *α_q_* satisfying the effectiveness of applied quarantine at the time *t_qc_* (Figure 3). *T_q,peak_* = *t_q,peak_* − *t_q_* is the time period during which *α* crosses line *α =* 1 and the number of new infection cases *n_I_* reaches maximum value after the quarantine start *t_q_*. During the next stage of constant growth rate *α = α_q_* < 1, the epidemic is finally suppressing until the end of the epidemic at the time *t_q_*,*_end_*, after which only small number of new accidental infection cases are registered.

**Figure 3.**
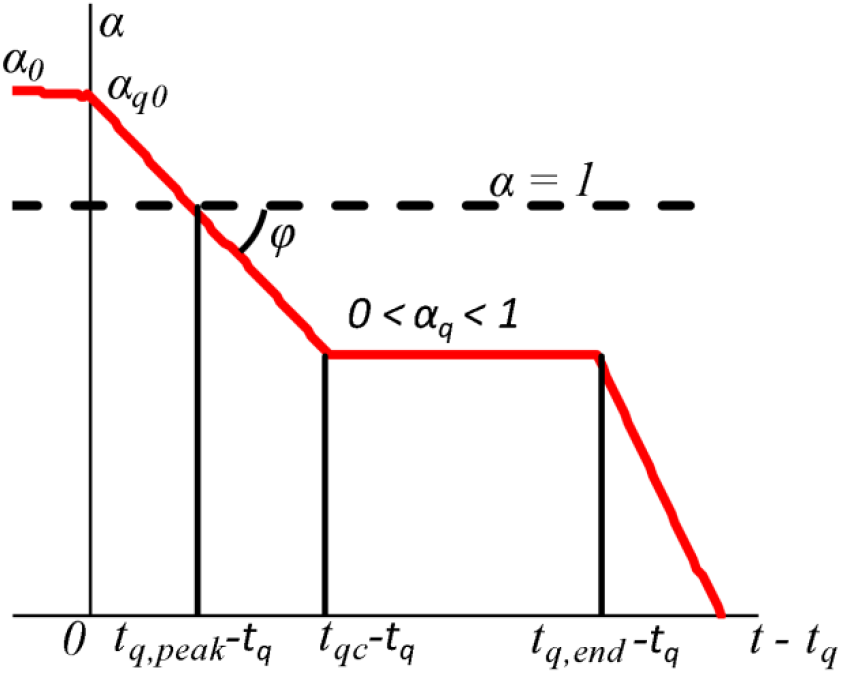
Realistic scenario of the growth rate of new cases *α* dynamics.

Duration of α decreasing stage *t_qc_* − *t_q_* depends on the properties of Covid-19 disease, such as incubation period and time span of individual being infectious, and how quickly quarantine actions are implemented. As a result, the angle *φ* = − *arctg* ((*α_q_*_0_ − *α_q_*)/(*t_qc_* − *t_q_*)) depends on *t_qc_* − *t_q_* and the effectiveness of applied quarantine actions, because the case of *φ* = 0 corresponds to situation when applied measures does not improve quarantine effectiveness and *e_q_* remains constant. The case of *φ* approaching to 90^0^ demonstrates exceptional effectiveness of the applied measures. Negative *φ* corresponds to increasing the growth rate of new cases *α*, what signalizes about critical state of broken quarantine. In addition, for a big country with heterogeneous population distribution across the country, it is expected to have several infection clusters and, therefore, wider time period *t_qc_* − *t_q_* will be for *α* decreasing stage (and consequently sharper *φ*) due to different start time of disease in each infection cluster. So, angle *φ* depends on the quarantine effectiveness, durations specific for the disease and homogeneity of the infected country or region.

The proposed parameters *e_q_* and *φ* together with analysis of the population mobility and social contacts [20] can be used to estimate effectiveness of country or region lockdown measures. In this paper we will analyse only *e_q_*.

In order to predict Covid-19 disease spread in infected country or region with imposed quarantine, a model of the growth rate of new cases *α* needs to be developed. It is possible to build up such model speculatively in general; however, it is reasonable to analyse dynamics of *α* in various countries. We analysed Covid-19 pandemic data from various countries [2,17]. The growth rate of new infection cases *α* was estimated on the basis of the registered daily new infection cases *n_I,d_*(*t*) (defined by eq. (7)) and smoothed by moving average according to eq. (9) with sliding window of length *N_s_* = 7 days in order to damp daily and weekly fluctuations:

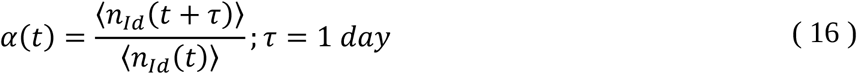

In this case, the parameter *α*(*t*) can be interpreted as the average daily reproduction number *R_t_* [19]. As illustration, the dynamics of 〈*α*〉, obtained as the parameter *α* smoothed by moving average according to eq. (9) with sliding window of length *N_s_* = 7 days, is demonstrated in Figure 4 for 53 countries (Albania, Algeria, Argentina, Australia, Austria, Azerbaijan, Bangladesh, Belarus, Belgium, Bosnia and Herzegovina, Chile, China, Croatia, Denmark, Ecuador, Estonia, Finland, France, Germany, Greece, Hungary, Iceland, Ireland, Italy, Japan, Kazakhstan, Latvia, Lithuania, Luxembourg, Malaysia, Malta, Moldova, Montenegro, Netherlands, New Zealand, Norway, Poland, Portugal, Romania, Russia, San Marino, Serbia, Singapore, Slovakia, South Korea, Spain, Sweden, Switzerland, Thailand, Tunisia, Ukraine, United Kingdom, United States of America) with imposed quarantine. The applied control measures of the quarantine in each country are described in [16]. Time is aligned to the date of imposed quarantine in each country *t*′ = *t* − *t_q_*, where *t_q_* is the time of start of the imposed quarantine [16]. Daily new infection cases until 20 May 2020 were provided by European Centre for Disease Prevention and Control [2].

**Figure 4.**
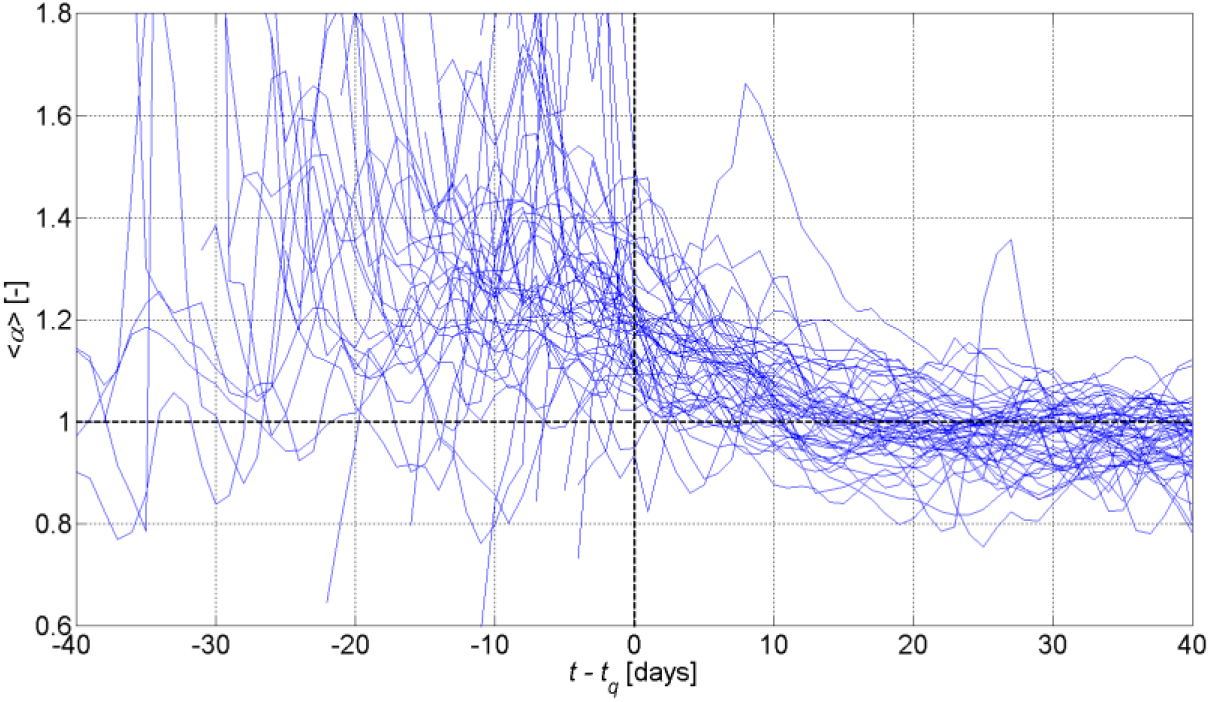
The averaged growth rate of new infection cases 〈*α*〉 versus time for 46 countries. Vertical black line points the day of imposed quarantine.

As can be seen, that despite of *α* fluctuations, the scenario described in Figure 3 can be identified in the dynamics of the parameter *α*: chaotic behaviour of *α* before quarantine demonstrates *α* linear decreasing until some certain value after quarantine. Let us analyse some examples in greater detail.

The typical example of above described scenario was realised in **Australia**, in which the epidemic started on 2020 January 26, the quarantine was imposed on 23 March 2020 [16] and epidemic is almost finished on May 03, 2020 with total number 6783 of confirmed infected cases. The dynamics of the number of daily new infection cases *n_Id_* [2] is presented in Figure 5 (a) by solid blue line and the moving average (with sliding window of length *N_s_* = 7 days) of daily new infection cases 〈*n_Id_*〉 by solid green line. The growth rate of new cases *α* estimated by eq. (16) is presented in Figure 5 (b) by solid blue line.

**Figure 5.**
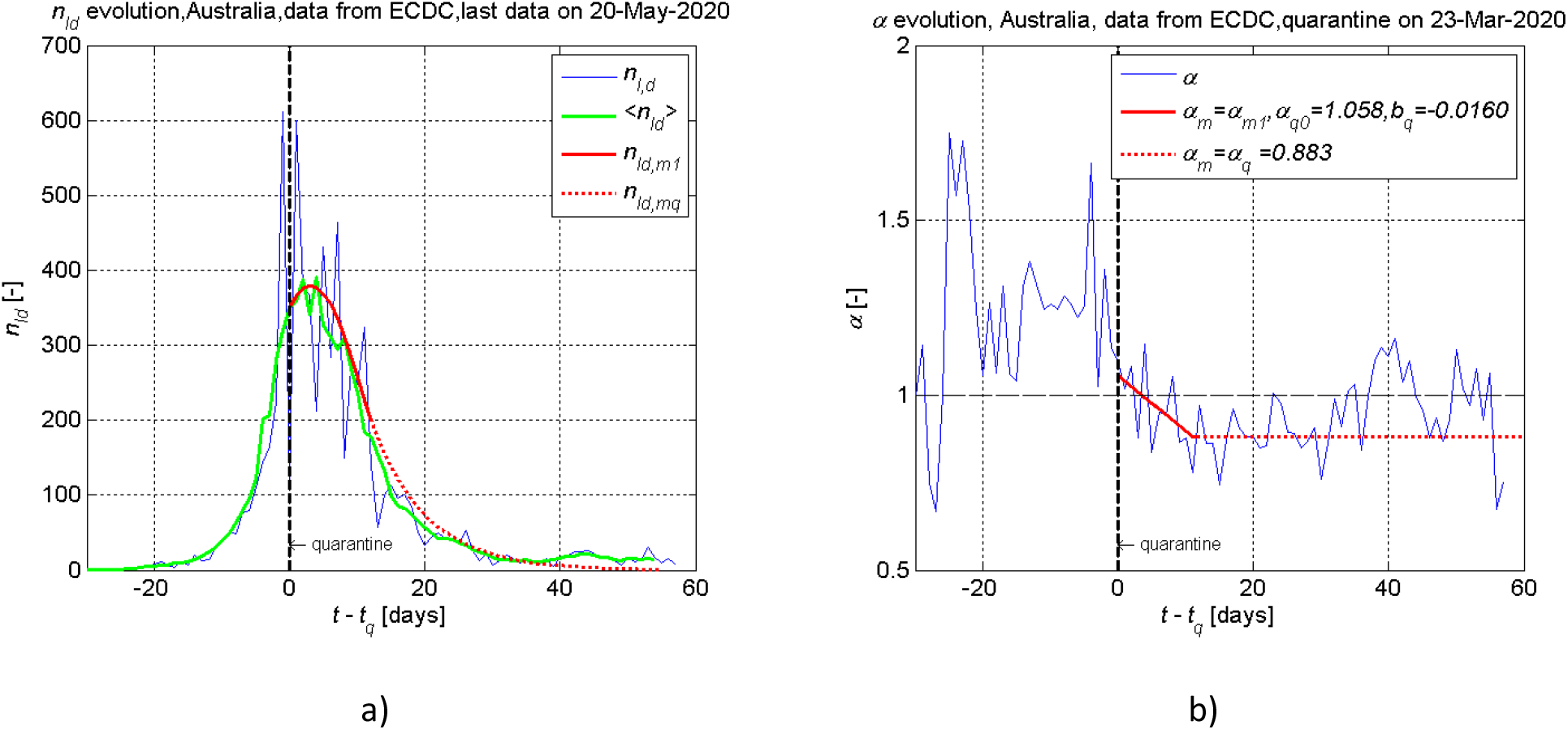
Dynamics of the daily new cases *n_Id_* (a) and the growth rate of new cases *α* (b) for Australia. Vertical black line points the day of imposed quarantine on 2020 March 23. Data collected from [2].

If to average fluctuations over supposed straights, the growth rate of new cases *α* was almost constant or slowly decreasing before quarantine, started to decrease sufficiently after quarantine had been imposed and had somewhat linear dependency on time during quarantine time, which allowed to approximate *α* by descending straight line (Figure 5 (b)) during the time period *t_qc_*− *t_q_ =* 12 days, where *t_qc_* is the date of α becoming equal to *α_q_* = 0.883. *α_q_* was estimated as mean value of *α* in time span of *t′* = [13,30] days, because after *t′* = 3 0 days only small numbers of daily new infection cases were registered because of casual nature of new infections. Then *α* was extended as constant until the epidemic end

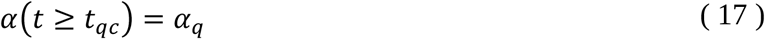

Consequently, the model of the growth rate of new cases can be described as follows:

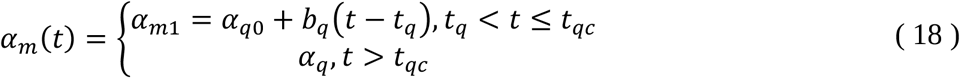

where *α*_*q*0_ = 1.058 is *α* value in the beginning of the quarantine

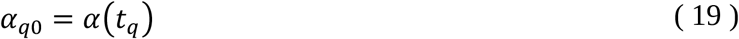

the parameter *b_q_* = −0.0160 characterizes decreasing of *α_m_* during the quarantine

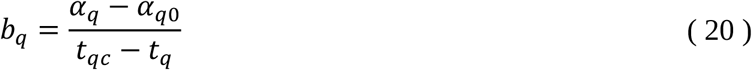

The modelled daily new infection cases *n_Idm_* is calculated by the simple algorithm

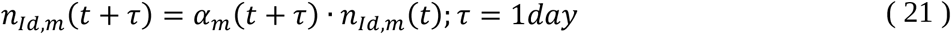

starting from the day of imposed quarantine *t* = *t_q_*: *n_Id,m_*(*t_q_*) = 〈*n_Id_*(*t_q_*)〉. The dynamics of *n_Id,m_*_1_ estimated by (*α_m_*_1_) for the time period *t_q_* < *t* ≤ *t_qc_* is shown by red solid line and *n_Id,mq_* estimated by *α_q_* for the time *t* > *t_qc_* by dashed red line.

The initial value of the growth rate of new cases can be estimated approximately as *α*_0_ ≈ 1.24. Consequently according to eq. (15), the estimated quarantine effectiveness is equal to *e_q_* = 1 − *α_q_*/*α*_0_ ≈ 0.29 or 29 %.

Occasional new infections occurring during last 4 weeks demonstrate that efficiency of applied quarantine actions was sufficient to suppress substantially the epidemic in the country, but the effectiveness is still not enough to decimate the infection.

Let us note, that small numbers of daily new infection cases (*n_Id_* < 2 0) are registered after *t′* = 30 days, and therefore, the proposed method cannot not be used for such small numbers, because of too casual nature of new infections. Consequently, the epidemic end cannot be forecasted accurately.

We will shortly overview some other countries demonstrating applicability and possible shortcomings of the proposed approach.

The next example of the similar scenario is **Switzerland**. The government of Switzerland announced that no lockdown would be implemented; however, some restrictions were implemented [21]. On 13 March 2020, the Federal Council decided to cancel classes in all educational establishments until 4 April 2020 and banned all events (public or private) involving more than 100 people. Furthermore, the borders were closed, and border control was enacted. On 16 March 2020, the Federal Council announced further measures and a revised ordinance. Measures included the closure of bars, shops and other gathering places until 19 April 2020, but leaved open certain essentials, such as grocery shops, pharmacies, (a reduced) public transport and the postal service. Since 20 March 2020, all events or meetings over 5 people were prohibited, and economic activities would continue including construction.

The growth rate of new cases *α* began decreased before first official restrictions of social contacts (Figure 6 (b)) while daily numbers of new cases were relatively small. During three following weeks after the restrictions initiation, *α* continued to decrease from 1.207 on 13 March 2020 until 0.927 and it is expected for *α* to remain constant until the end of the epidemic (Figure 6).

**Figure 6.**
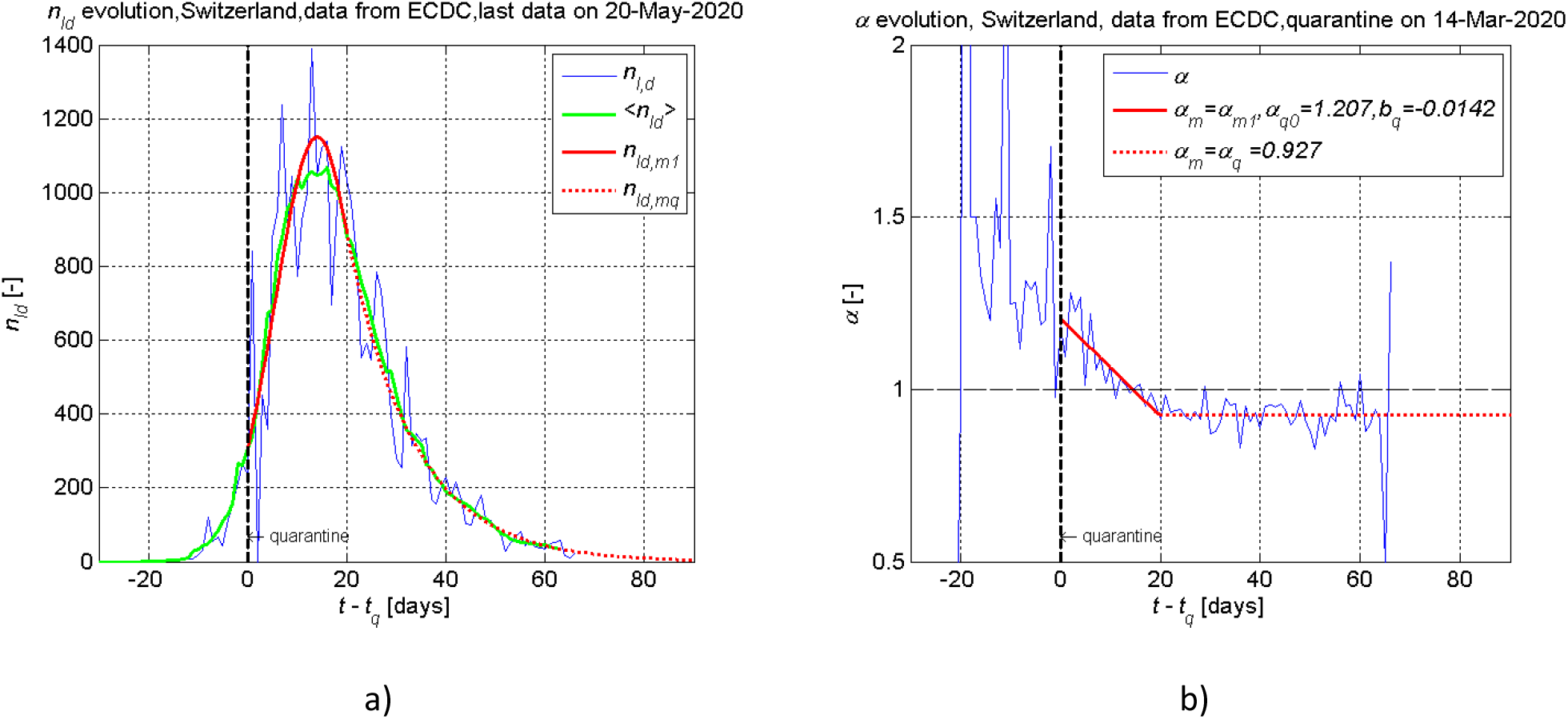
Dynamics of the daily new cases *n_Id_* (a) and the growth rate of new cases *α* (b) for Switzerland. Vertical black line points the day of first quarantine action on 2020 March 13. Data collected from [2].

**New Zealand** is an example of slightly different scenario (Figure 7). On 21 March, 2020 at midday, New Zealand Prime Minister announced the introduction of a country-wide alert level system, similar to the existing fire warning systems [22]. There are four levels, with 1 being the least risk of infection and 4 the highest. At the time of Prime Minister’s announcement she stated that New Zealand was at level 2. Each level brings added restrictions on activities or movements. Prime Minister announced on 23 March at around 2 pm that, effective immediately, New Zealand would be at alert level 3, moving to level 4 at 11:59 pm on 25 March.

**Figure 7.**
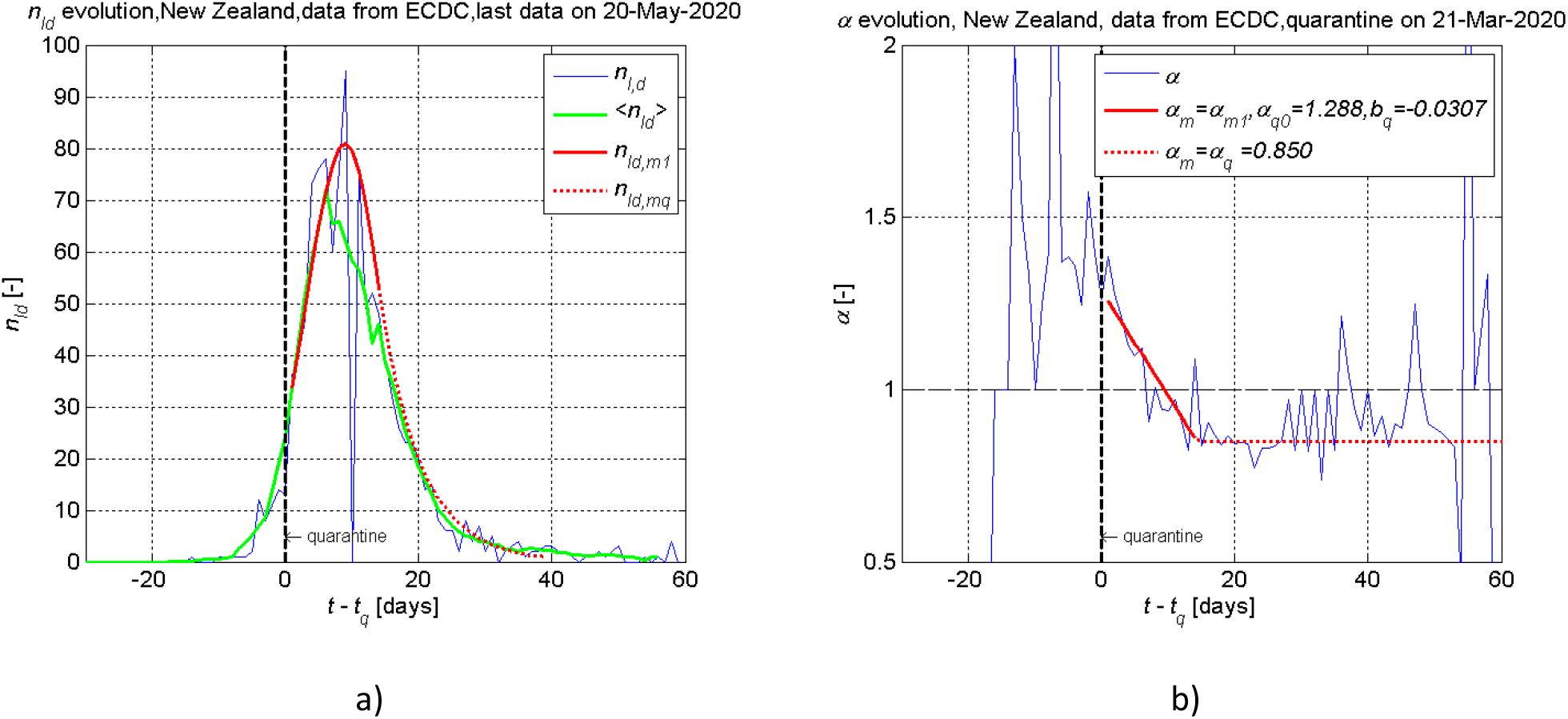
Dynamics of the daily new cases *n_Id_* (a) and the growth rate of new cases *α* (b) for New Zealand. Vertical black line points the day of imposed quarantine on 2020 March 21. Data collected from [2].

The dynamics of the daily new infection cases *n_Id_* [2] and the calculated growth rate of new cases are presented in Figure 7. The modelled *n_Id_* was overestimated (Figure 7 (b)), because *α* was approximated by one straight in the time interval from *t_q_* (corresponding to 21 March 2020) to *t_qc_* = *t_q_* + 14 days (Figure 7 (a)). To improve the model of *α*, the change of alert level from 2 to 4 on fifth day after the beginning of the quarantine must be considered. Accordingly, *α* must be approximated by 2 straights in the time interval [*t_q_*,*t_qc_*].

The similar scenario is demonstrated in **Iceland**, where universities and secondary schools were closed on 16 March 2020. Furthermore, public gatherings of over 100 were banned on the same day [23]. The same quarantine conditions were applied for the whole quarantine period. Nevertheless, there were three stages during the time interval [*t_q_*,*t_qc_*], *t_qc_ = t_q_ +* 25 days (Figure 8 (b)). The first stage of decreasing *α* lasted for 6 days since the beginning of the quarantine. Then the next stage followed up for 7 days, in which the *α* fluctuates around *α ≈* 1. The third and the final stage of decreasing *α* took place for 12 days, during which *α* reached value of *α_q_ =* 0.863. Therefore, *α* model for the whole period of 25 days, which describes the behaviour of *α* by one decreasing line *α_m_*_1_ = *α_q_*_0_ + *b_q_* (*t* − *t_q_*), *t_q_* < *t* < *t_qc_* overestimates the number of daily new infection cases *n_Id_* in the second stage (Figure 8 (a)).

**Figure 8.**
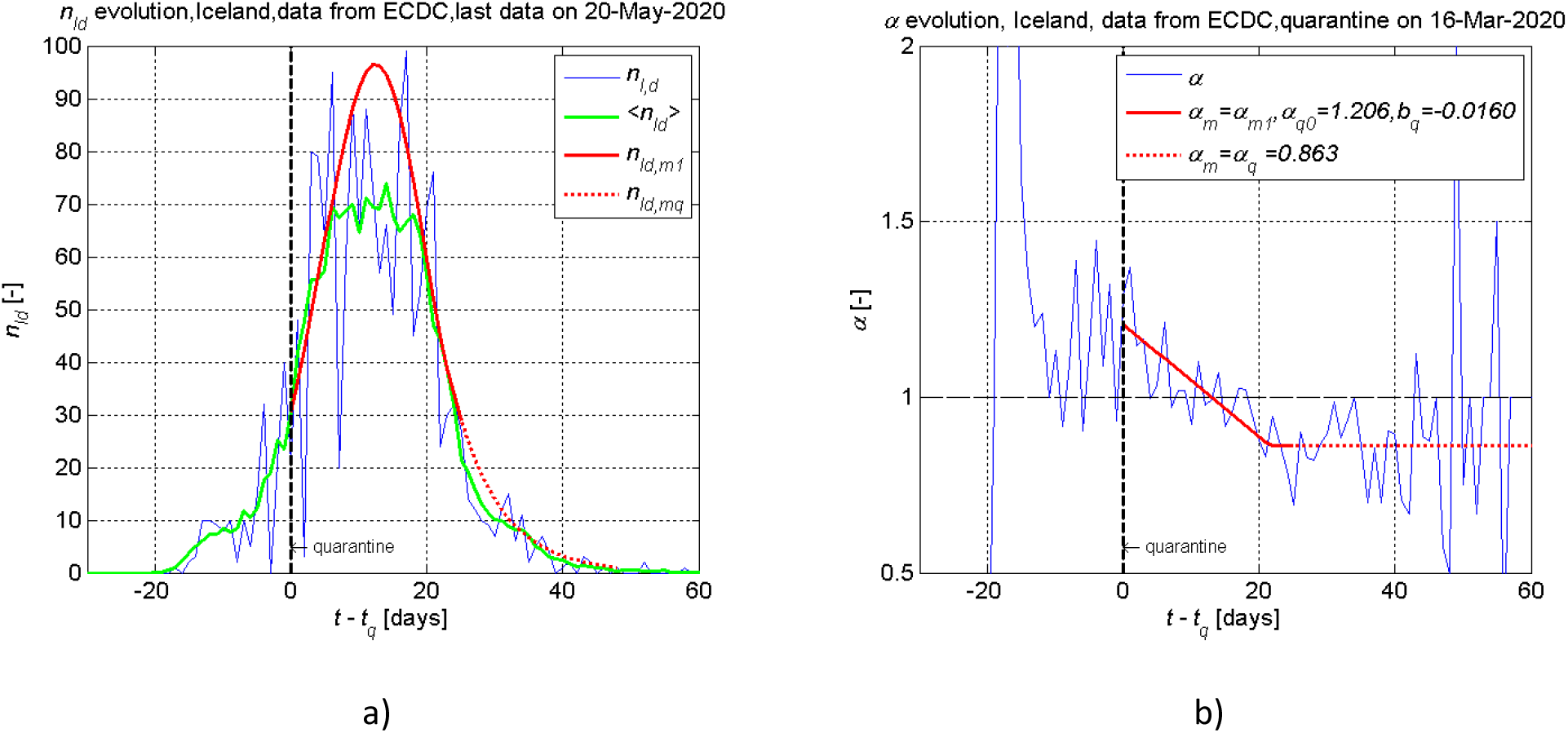
Dynamics of the daily new cases *n_Id_*; (a) and the growth rate of new cases *α* (b) for Iceland. Vertical black line points the day of imposed quarantine on 2020 March 16. Data collected from [2].

The similar scenario to Australia and Switzerland cases developed in **Austria** (Figure 9), where the epidemic started on 2020 February 25, 2020 and the quarantine was imposed on 2020 March 16 [24]. However, slight increase of the growth rate *α* after one month of quarantine may be related to the Easter celebration, which started after Good Friday, 2020 April 10. After the Easter, the number of daily new cases fluctuates around 65 cases per day.

**Figure 9.**
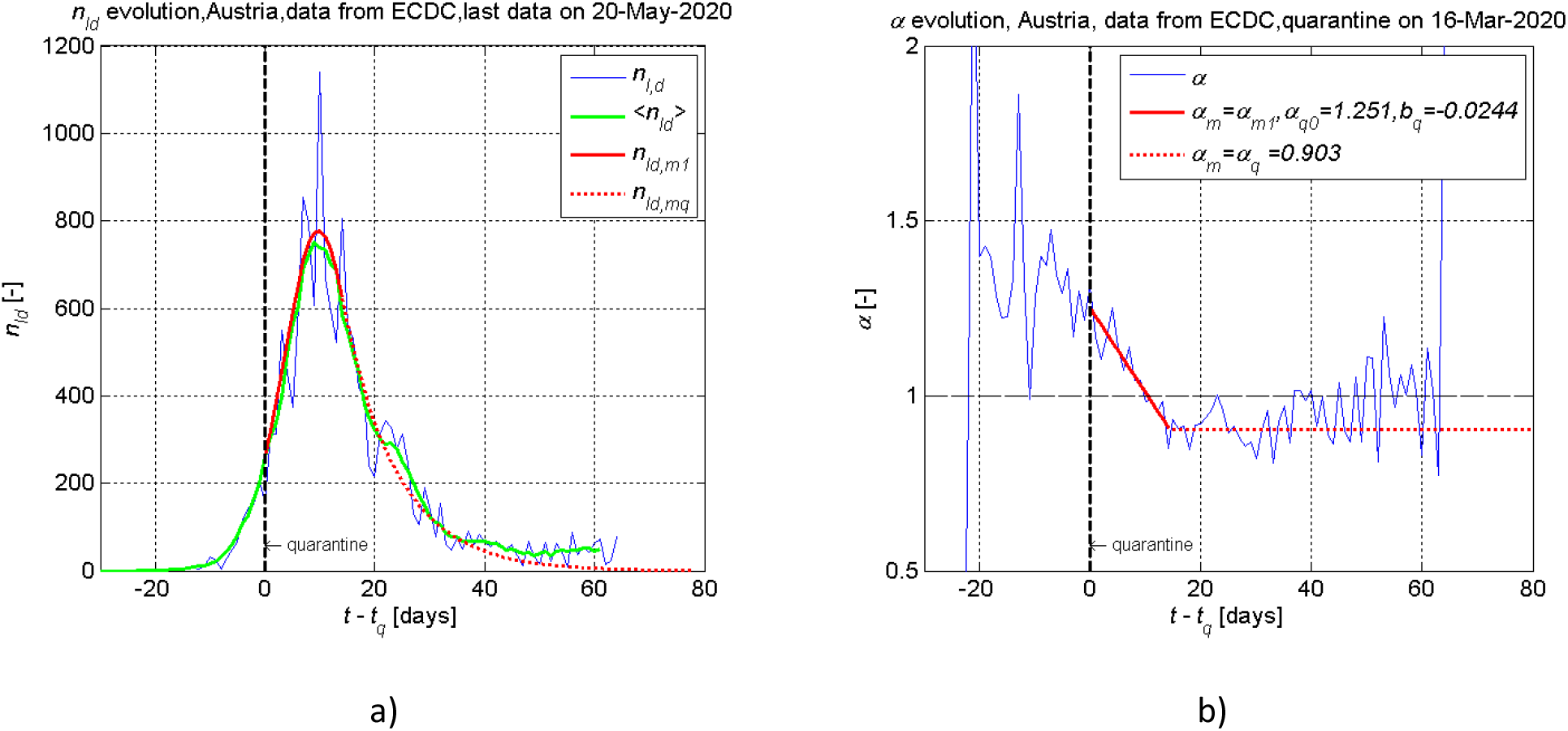
Dynamics of the daily new cases *n_Id_* (a) and the growth rate of new cases *α* (b) for Austria. Vertical black line points the day of first quarantine action on 2020 March 16. Data collected from [2].

The case of **Poland** illustrates the scenario when the proposed method to predict epidemic dynamics under quarantine does not work straightforward, because of country specificity. Strict rules of the country lockdown were implemented on 12 March 2020 [21], and it was expected, that quarantine effectiveness would be the same as in the cases described above. However, during four following weeks after the beginning of the quarantine, *α* decreased from 1.256 only to 1.0 and then stayed fluctuating around *α* ≈ 1 till now (20 May 2020) (Figure 10), what shows the low effectiveness of the taken quarantine actions. As in Austria case, the most reliable reason is the Easter celebration. During the expected next wave of the Covid-19, such specificity of the country can be taken in to account.

**Figure 10.**
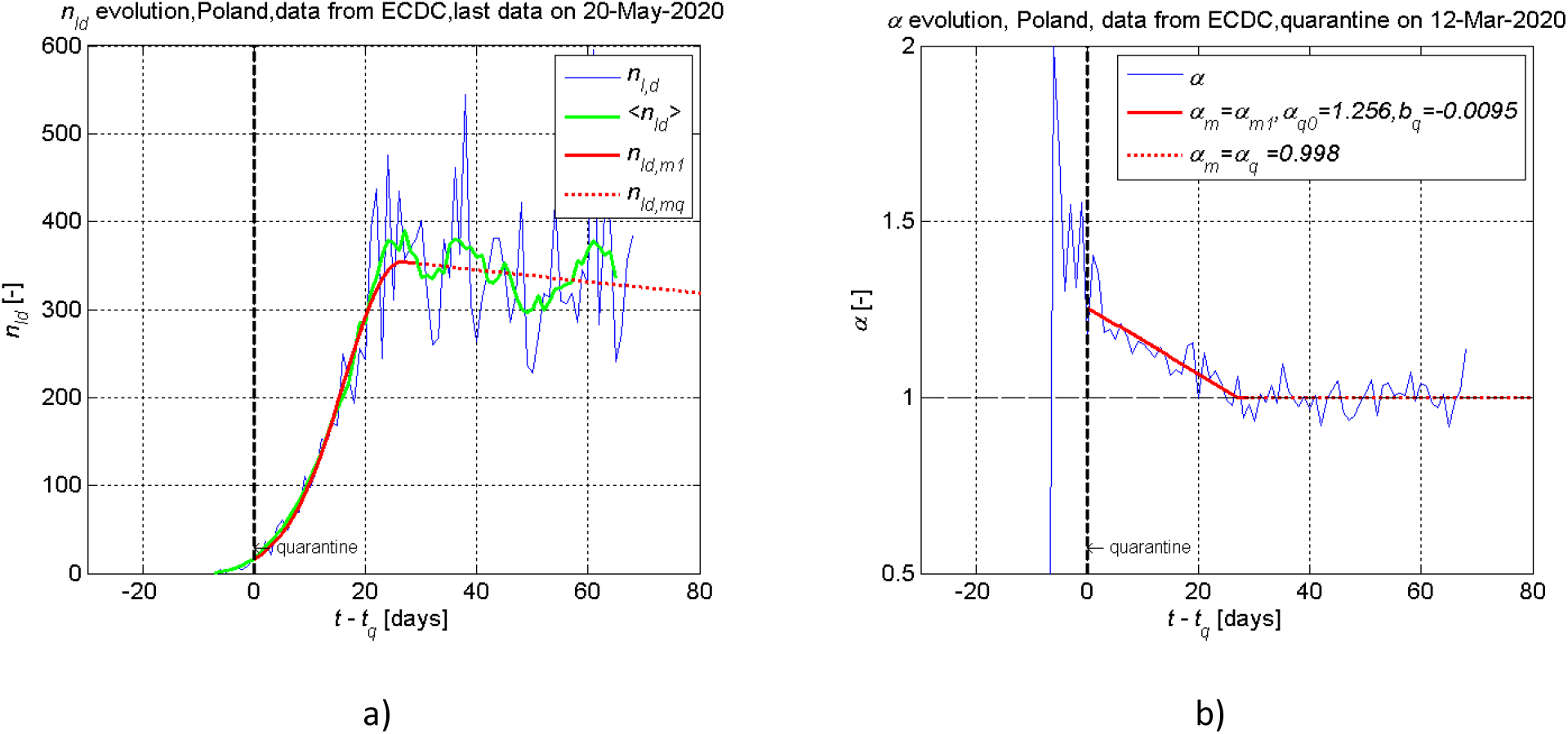
Dynamics of the daily new cases *n_Id_* (a) and the growth rate of new cases *α* (b) for Poland. Vertical black line points the day of imposed quarantine on 2020 March 12. Data collected from [2].

**Italy** has been under quarantine since March 9, 2020 [16]. It seems that Italians are too tired of the country lockdown, that they have strong intentions to finish the lock-down as soon as possible. Therefore, Italy celebrated the Easter more quietly without visible breaks of the quarantine restrictions (Figure 11). However, a high value of the growth rate of new infection cases during the imposed quarantine *α_q_* = 0.960 suggests that quarantine actions are not sufficient and, there is little hope to reach the end of epidemic as fast as Australia having *α_q_* = 0.883.

**Figure 11.**
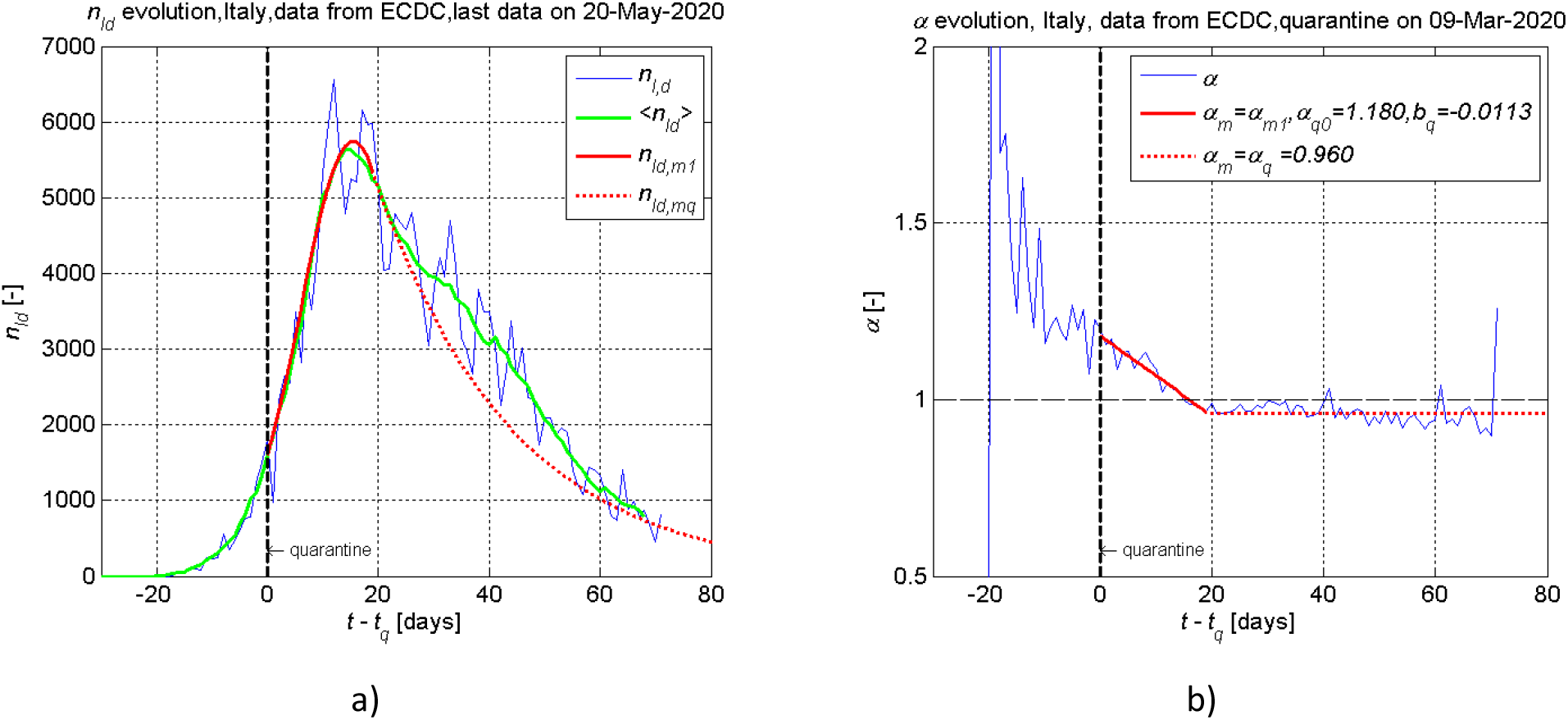
Dynamics of the daily new cases *n_Id_* (a) and the growth rate of new cases *α* (b) for Italy. Vertical black line points the day of imposed quarantine on 2020 March 09. Data collected from [2].

The evolution of *α* shows that the Easter celebration had no substantial influence on quarantine effectiveness in **Germany** (Figure 12), which is under strict national quarantine since 23 March 2020 [25].

**Figure 12.**
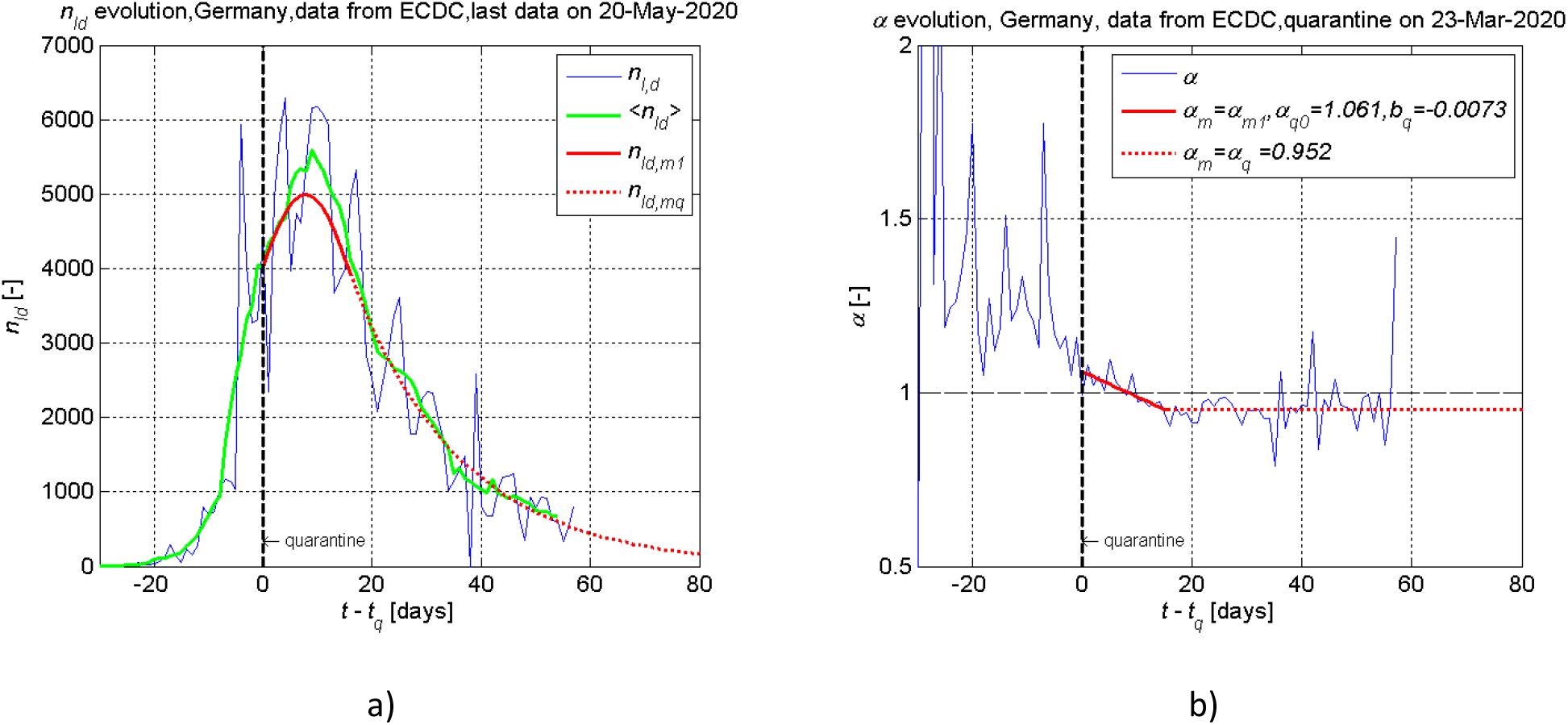
Dynamics of the daily new cases *n_Id_* (a) and the growth rate of new cases *α* (b) for Germany. Vertical black line points the day of imposed quarantine on 2020 March 23. Data collected from [2].

**The United Kingdom** imposed the national quarantine on 24 March 2020 [25] and reached a peak of daily new infection cases in three weeks later (Figure 13). However, as in Italy case, a high value of the growth rate of new infection cases during the imposed quarantine *α*_*q*_ = 0.983 suggests that quarantine actions are not sufficient to reach the end of the epidemic as fast as in Australia.

**Figure 13.**
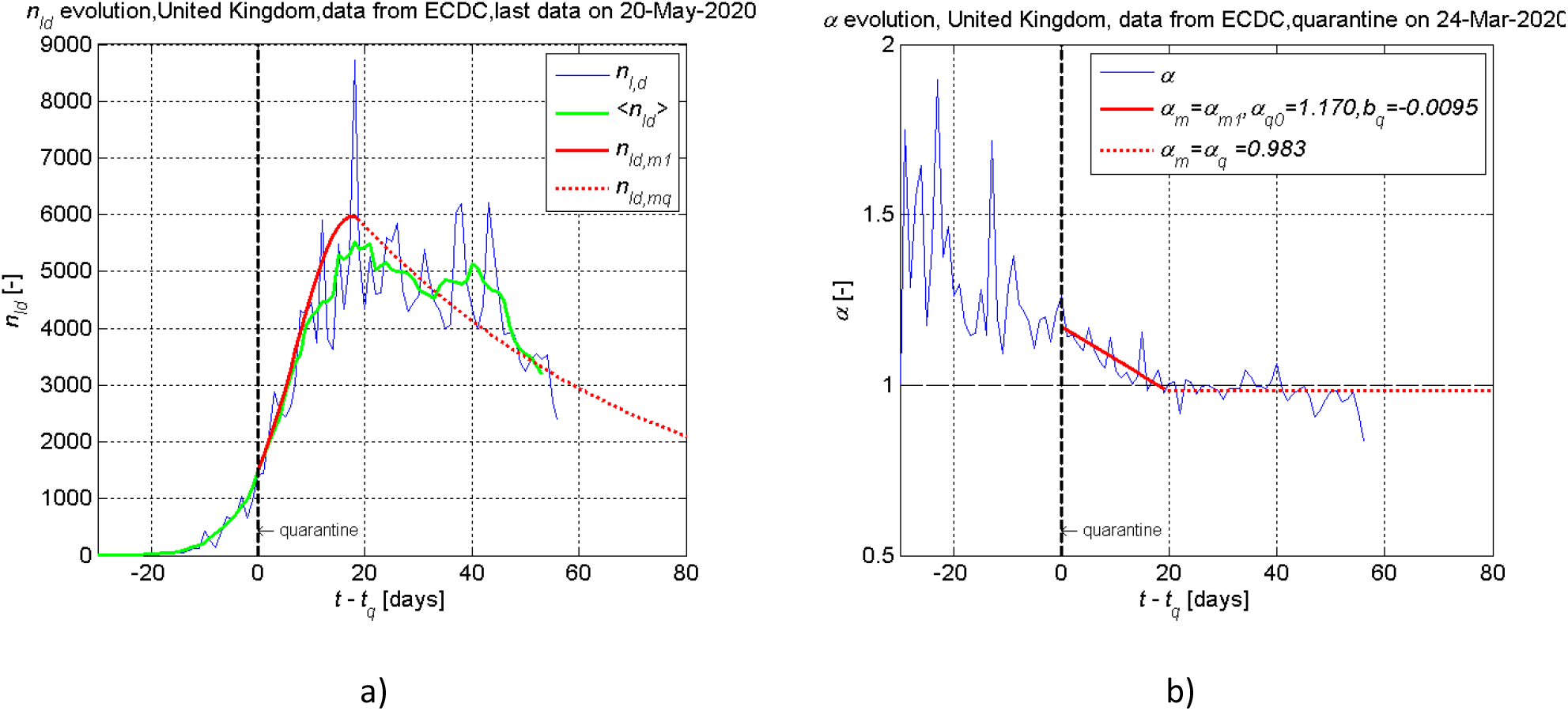
Dynamics of the daily new cases *n_Id_* (a) and the growth rate of new cases *α* (b) for the United Kingdom. Vertical black line points the day of imposed quarantine on 2020 March 24. Data collected from [2].

**Denmark** is an example in which the proposed model cannot be applied to predict the epidemic dynamics, and clarification of the reasons for such discrepancy requires more detailed analysis of epidemic situation in the country (Figure 14). However, the last 35 days *α* is below 1, what demonstrates that social distancing helps to reduce infection rate.

**Figure 14.**
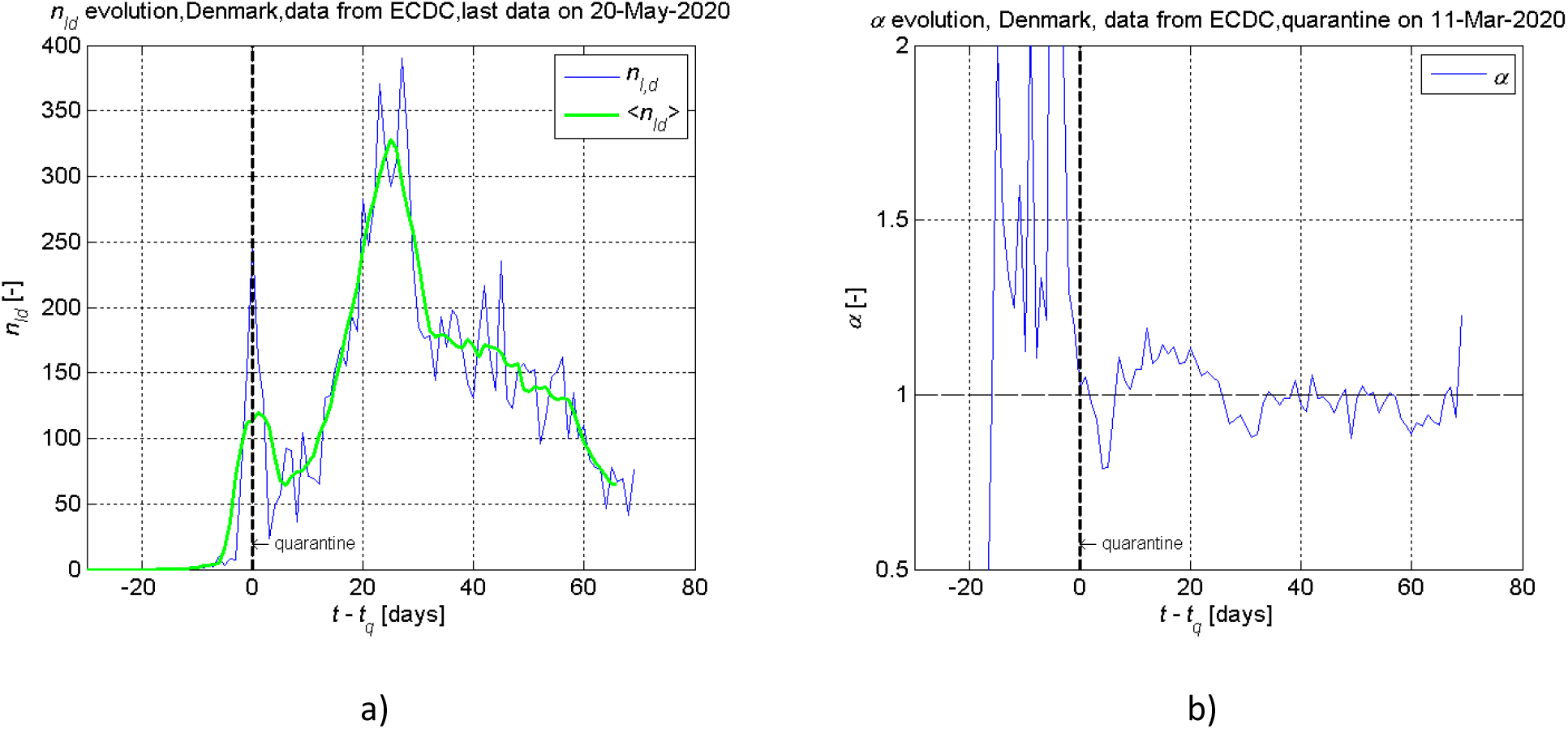
Dynamics of the daily new cases *n_Id_* (a) and the growth rate of new cases *α* (b) for Denmark. Vertical black line points the day of imposed quarantine on 2020 March 11. Data collected from [2].

Situation in **Russia** serves as an example of ineffective quarantine actions for at least first two weeks of the country lockdown. This can be explained by huge size of the country and heterogeneous distribution of the population across the country, which is reason for sequence of arising infection clusters in different locations/regions at different times, despite that national quarantine was imposed on 30 March, 2020. Consequently, the growth rate of new infection cases *α =* 1.146 remained above 1 for 18 days with almost zero angle of inclination *φ =* − *arctg*(*b_q_*) ≈ (Figure 15 (b)), what demonstrates that effectiveness of the quarantine actions was insufficient during this period. For example, only from 12 May, Moscow’s residents will be required to wear face masks and gloves in public transport and public places [26], while face masks in public places are one of the most important obstacles for virus spread [27]. Consequently, the growth rate *α* crossed critical value *α =* 1 only after 40 days from the national lockdown start, but it gives hope that eventually the epidemic reached peak of the daily new infection cases *n_Id_*. Because of heterogeneous distribution of the population, analysis and forecasting of epidemic situation must be done at region level to generate results with practical value.

**Figure 15.**
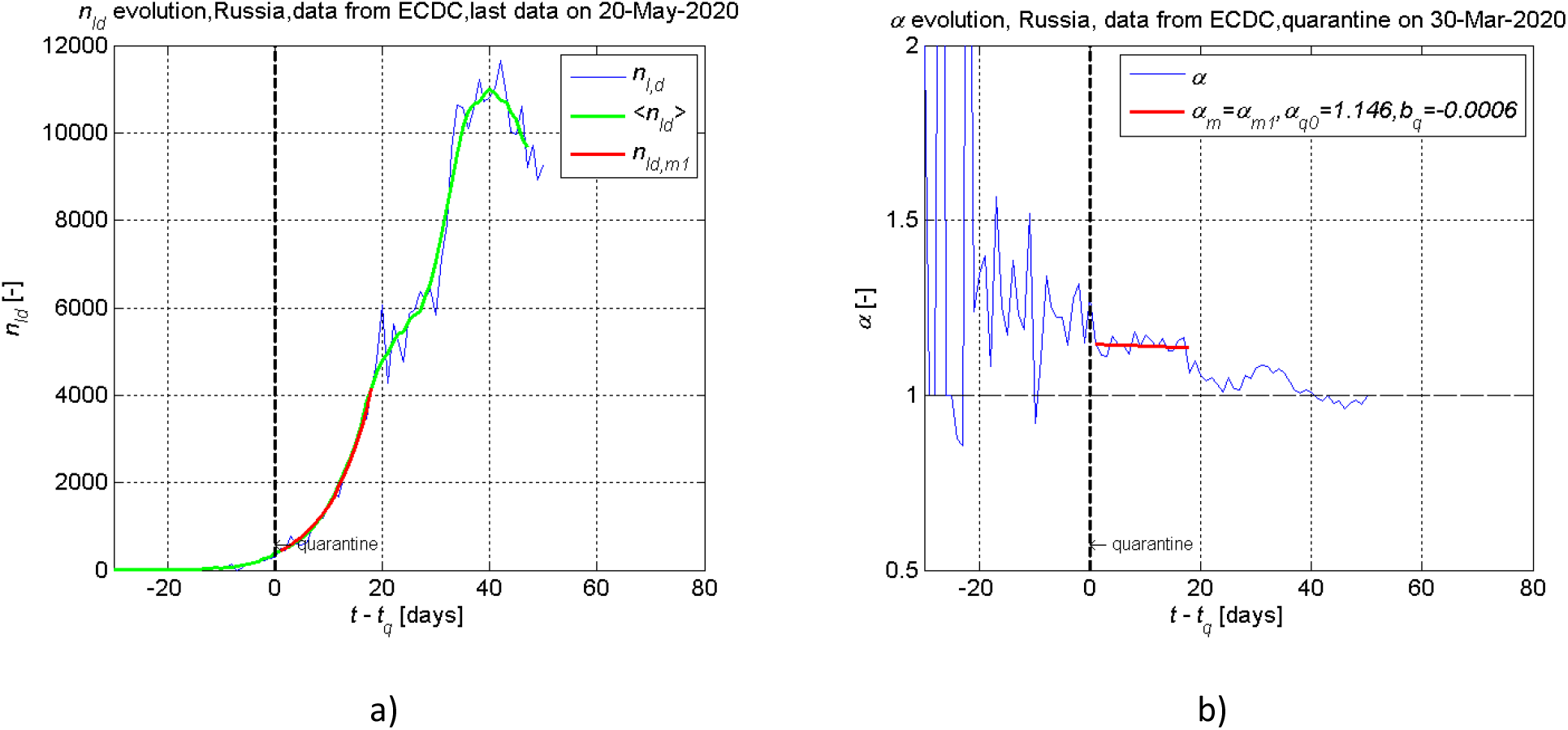
Dynamics of the daily new cases *n_Id_* (a) and the growth rate of new cases *α* (b) for Russia. Vertical black line points the day of imposed quarantine on 2020 March 30. Data collected from [2].

Like Russia, **United States of America** is another huge country by the population number and size. The dynamics of the new cases *n_Id_* and the growth rate of new infection cases *α* fluctuating around *a* ≈ 1 till now in USA show that quarantine is not effective enough (Figure 16) and suggested emergence of new clusters of infection. The forecast, made on the basis of the current dynamics of the epidemic, shows that epidemic will not be defeated in the USA until the end of this year. It should be taken into account, that differences of COVID-19 statistics across various states are huge: cases per 100 000 people are varying more than 10 times [28]. Therefore, overall US data can suffer from too high level of generalisation. Again as in Russia, analysis and forecasting of epidemic situation should be done at state level to generate results with practical value.

**Figure 16.**
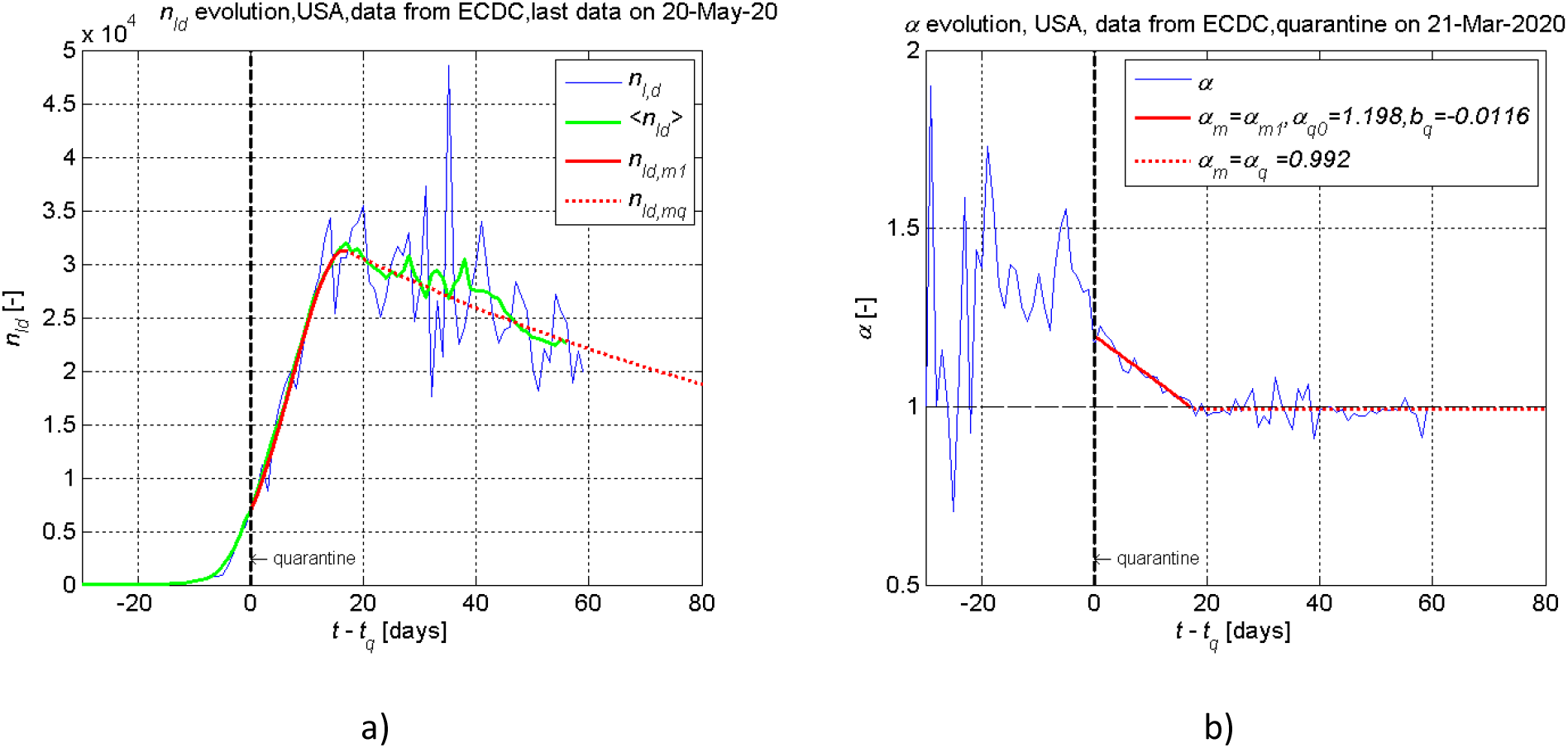
Dynamics of the daily new cases *n_Id_* (a) and the growth rate of new cases *α* (b) for USA. Vertical black line points the day of supposed average quarantine date 2020 March 20. Data collected from [2].

Scenario realised in **Sweden**, where the quarantine started on 10 March, 2020 [29], is specific. Due to soft conditions of the quarantine (technically most of EU countries would not attribute Swedish regime a quarantine, just a gradual restriction of some social activities), it seems that the new cases *n_Id_* is achieved maximum only in 6 weeks days after the lockdown start and then the *α* stabilised at *α_q_ =* 1 (Figure 17). Further dynamics of the epidemic is still unclear because no country has experience of such situation. The experience gained in Sweden is very important and will be used by other countries for the second wave management in the future.

**Figure 17.**
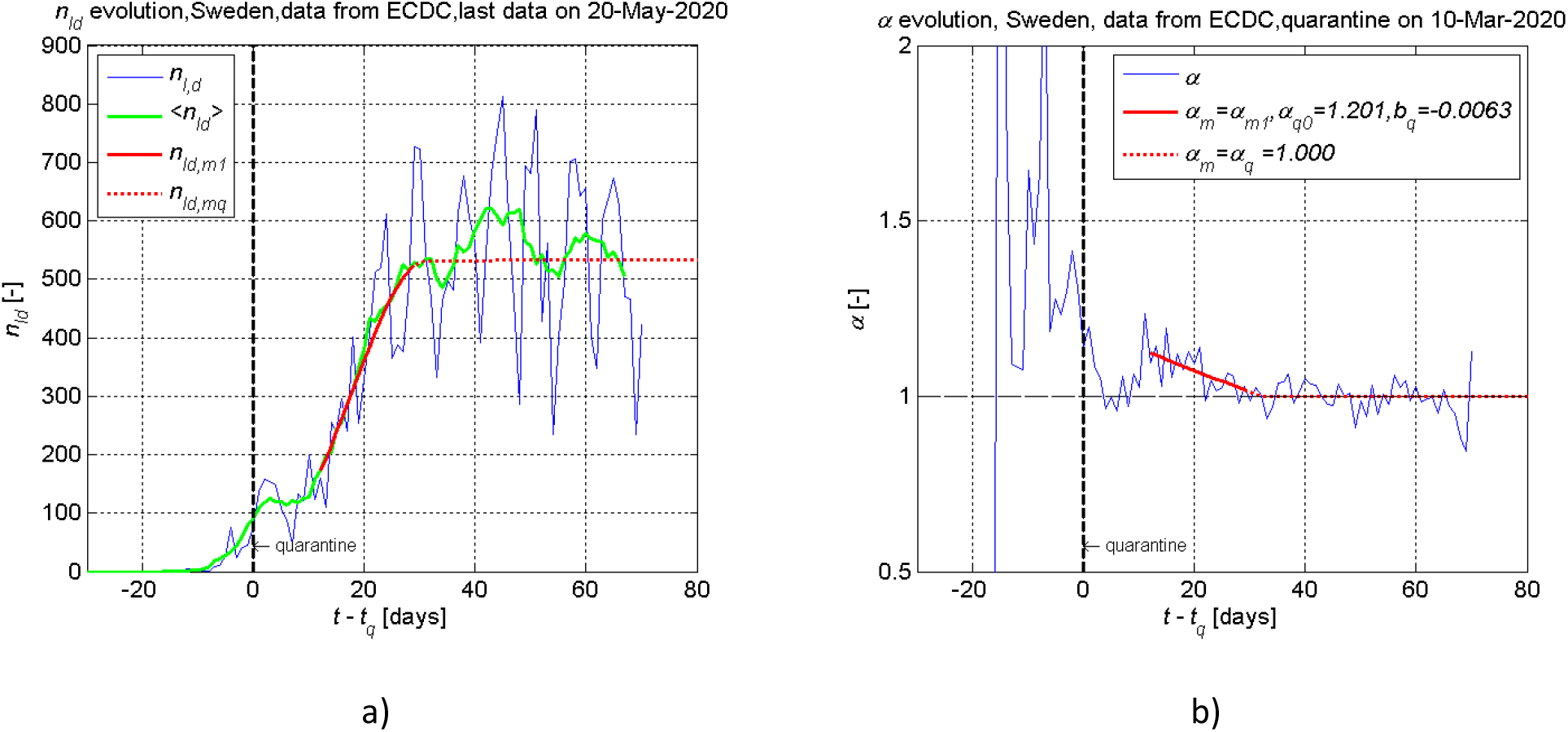
Dynamics of the daily new cases *n_Id_* (a) and the growth rate of new cases *α* (b) for Sweden. Vertical black line points the day of imposed initial quarantine actions on 2020 March 10. Data collected from [2].

## Conclusions

The experience gained during the first wave of Covid-19 pandemic could help countries to be better prepared for the next wave, which is expected to take place in the autumn of 2020. We proposed the simplified approach, which allows to quantifiable estimate effectiveness of the imposed quarantine conditions/restrictions and compare between countries, forecast the epidemic spread and take appropriate decisions. The proposed approach is based on the time series data of registered daily infection rate as most reliable epidemiological data together with the following main assumptions:

- Total number of infections is much smaller than a population size of the infected country or region;
- Registered infected individuals are effectively isolated during the imposed quarantine.

The observed dynamics of the pandemic in examined countries shows that the growth rate of new infection cases has a tendency to decrease linearly during the imposed quarantine period until reaching a constant value, which corresponds to the effectiveness of taken quarantine measures in the country. The proposed parameters *α_q_*, *e_q_* and *φ* together with the analysis of the population mobility and social contacts can be used to estimate the effectiveness of the lockdown measures. On the basis of these parameters, the countries experiencing ongoing epidemic can use the proposed approach to study effectiveness of taken quarantine measures in other countries yet affected by the Covid-19 disease.

The proposed approach has a limitation because it cannot be directly applied for a country with large population size, which might have several epidemic clusters due to the heterogeneous population distribution across the country. In this case, each cluster must be analysed separately.

Also, the approach cannot be applied when the daily infection cases is too few because of casual nature of emerging new infections. Consequently, the epidemic finish cannot be forecasted accurately.

On the basis of the proposed approach, more complex models of epidemic forecasting can be developed.

## Data Availability

The data used to support the findings of this study are available from the corresponding author upon request. Data for the infected cases count from various countries is obtained from the data collected in [2,17]. Dates of the imposed quarantine were collected from [16].

https://en.wikipedia.org/wiki/Template:2020_coronavirus_quarantines_outside_Hubei

https://www.ecdc.europa.eu/en/publications-data/download-todays-data-geographic-distribution-covid-19-cases-worldwide

https://data.humdata.org/dataset/novel-coronavirus-2019-ncov-cases

## Acknowledgements

This research is funded by the Research Council of Lithuania under the project P-MIP-17-108 “ComDetect” (Agreement No. S-MIP-17-69), 2017-2020.

We also would like to thank Tadas Markūnas and Aušra Džiugytė for collecting of Covid-19 epidemiological data.

## Authors’ contributions

AD developed the method, analysed data and post-process data. MB proposed the conception of the model and analysed data. GS, EM and RN performed data processing. All authors wrote and approved the manuscript.

## Additional information

**Correspondence and requests for materials** should be addressed to AD.

